# An automated heart rate-based algorithm for sleep stage classification: validation using conventional PSG and innovative wearable ECG device

**DOI:** 10.1101/2021.12.21.21268117

**Authors:** Nicolò Pini, Ju Lynn Ong, Gizem Yilmaz, Nicholas I. Y. N. Chee, Zhao Siting, Animesh Awasthi, Siddharth Biju, Kishan Kishan, Amiya Patanaik, William P. Fifer, Maristella Lucchini

**Affiliations:** Department of Psychiatry, Columbia University Irving Medical Center, New York, NY, USA; Centre for Sleep and Cognition, Yong Loo Lin School of Medicine, National University of Singapore, Singapore; Electronic and Information Engineering, Imperial College, London, UK; Department of Biotechnology, Indian Institute of Technology, Kharagpur, India; Neurobit Inc., New York, NY, USA

**Keywords:** Sleep Staging and Scoring, Automatic Sleep Staging, Wearables, Heart Rate Variability, Artificial Intelligence

## Abstract

**Study Objectives:** Validate a HR-based deep-learning algorithm for sleep staging named Neurobit-HRV (Neurobit Inc., New York, USA).

**Methods:** The algorithm can perform classification at 2-levels (Wake; Sleep), 3-levels (Wake; NREM; REM) or 4-levels (Wake; Light; Deep; REM) in 30-second epochs. The algorithm was validated using an open-source dataset of PSG recordings (Physionet CinC dataset, n=994 participants) and a proprietary dataset (Z3Pulse, n=52 participants), composed of HR recordings collected with a chest-worn, wireless sensor. A simultaneous PSG was collected using SOMNOtouch. We evaluated the performance of the models in both datasets using Accuracy (A), Cohen’s kappa (K), Sensitivity (SE), Specificity (SP).

**Results:** <u>CinC</u> - The highest value of accuracy was achieved by the 2-levels model (0.8797), while the 3-levels model obtained the best value of K (0.6025). The 4-levels model obtained the lowest SE (0.3812) and the highest SP (0.9744) for the classification of Deep sleep segments. AHI and biological sex did not affect sleep scoring, while a significant decrease of performance by age was reported across the models. <u>Z3Pulse</u> - The highest value of accuracy was achieved by the 2-levels model (0.8812), whereas the 3-levels model obtained the best value of K (0.611). For classification of the sleep states, the lowest SE (0.6163) and the highest SP (0.9606) were obtained for the classification of Deep sleep segment.

**Conclusions:** Results demonstrate the feasibility of accurate HR-based sleep staging. The combination of the illustrated sleep staging algorithm with an inexpensive HR device, provides a cost-effective and non-invasive solution easily deployable in the home.

## INTRODUCTION

Sleep is a biological necessity that leads humans to spend roughly one third of their life asleep. Emerging research is highlighting the critical role that sleep plays in overall health. Poor sleep health has been associated with several negative health outcomes, including increased risk of cardiovascular disease,^1,2^ obesity,^3^ depression,^4^ and neurodegenerative disorders.^5^ Despite these well-known health risks, we are in the middle of a sleep crisis, with more than 70 million Americans experiencing sleep related problems^6^ and with less than 20% of patients estimated to be properly diagnosed and treated for sleep disorders.^7^

Polysomnography (PSG) is presently considered the gold standard method to assess sleep and is performed in sleep laboratories and clinical settings. It records simultaneously electroencephalogram (EEG), electromyogram, electroocular activity, heart rate, respiration, leg movements, nasal pressure, oxygen desaturation and body position and utilized these signals to characterize sleep architecture and sleep disorders. Limitations of PSG include the cost of the devices, laborious setup procedures, the discomfort, and the necessity to have an expert perform the time-consuming process of coding the data. These limitations highlight the need of noninvasive, inexpensive, and reliable sleep monitors that could be deployed in the home to monitor sleep health as well as automated solutions to perform reliable sleep stage scoring. While accelerometer-based actigraphy devices have been used in the field for decades, estimation is limited to sleep/wake states which is insufficient for a full evaluation of sleep architecture.

Recent technological advancements have led to a proliferation of consumer-oriented tools to monitor sleep.^8^ Initially these tools relied mainly on activity, similarly to actigraphy, but lately they have started to incorporate additional physiological signals, such as EEG, heart rate (HR), breathing and pulse oximetry.^9–11^ Increasing research indicates that HR, defined as the number of heart beats in 1 minute, and its variability might be an accurate and accessible physiological proxy for sleep measurement.^12–15^ A wealth of literature has shown profound differences in HR across sleep stages,^16–18^ primarily comparing REM versus NREM sleep. In adults, REM sleep has been reported to be associated with an increment in low frequency power of the HR variability signal, and a decrement in high frequency power.

Heart rate can be derived from electrocardiography (ECG), which records the heart’s electrical activity, or photoplethysmography (PPG), which monitors changes in blood volume in a portion of the peripheral microvasculature, estimating pulse waveforms. While PPG is more commonly employed in wearable devices^19,20^ because it can easily estimate HR from various peripheral body locations, several concerns have been raised regarding its reliability given the strong dependence on ambient light, skin conditions and colors, and dealing with physical motion artifacts.^21,22^ Thus, ECG-derived HR is still considered the gold standard, but it rarely used in wearable technologies. In addition, very often algorithms that utilize activity and physiological data to characterize sleep are proprietary and not fully/only partially validated against gold standard PSG measures. In most cases, raw data access is either unavailable or limited,^23,24^ and algorithm or firmware upgrades are also not always transparent to users, limiting longer-term comparisons either within or between individuals. In summary, the ‘black-box’ nature of these devices and the paired firmware/software packages limits the application and deployment of these devices for clinical and research purposes.

To address this gap in the literature, Neurobit HRV (Neurobit Inc., New York, NY, USA) was designed, a sleep staging algorithm which utilizes HR parameters derived from a single-channel ECG. In this manuscript, we will show the performance of this algorithms on a public dataset (Physionet CinC^25^), and on a secondary dataset with data simultaneously acquired with PSG and an ECG patch. Results demonstrate the feasibility of accurate sleep staging based of HR derived from ECG, obtained from PSG or wearable devices. The combination of the proposed sleep staging algorithm with inexpensive and commonly used ECG-patches (∼100$), provides a cost-effective and non-invasive solution easily deployable in the home for large-scale sleep characterization in the field.

## MATERIAL AND METHODS

### The sleep staging algorithm

The HR based automated sleep staging software called Neurobit-HRV was developed by Neurobit Inc., New York, NY, USA. The software is open source and publicly available through the cloud as a software development kit (SDK, https://bitbucket.org/NeurobitTech/pyndf) as well as an application programming interface (API) along with reference code and documentation.^26^ The deep-learning algorithm was implemented in Python 3.6 using the Keras (https://keras.io/) and Tensorflow 2 library (https://www.tensorflow.org/). The algorithm was trained on private datasets comprising of ECG extracted from 12,404 PSG records primarily collected from academic sleep centers in South-East Asia (35%), North America (30%) and Europe (30%). Fifty-nine percent of the assessed participants had a suspected sleep disorder whereas the remaining 41% were from healthy subjects. The mean age of such aggregated dataset was (mean ± std) 42.3 ± 16.8 years. The observations were first randomly split into training (80%) and testing (20%) data. The split was stratified by the data source. The model which obtained the smallest test error was selected as the optimal one. Figure 1 displays the trends of model accuracy for the training and testing sets as a function of progressive iterations. The software can operate on either a single channel ECG or directly on R-peak locations. To achieve optimal performance, the temporal precision of the R-peak location must be ±4ms or better. Neurobit-HRV incorporates an extensive wavelet-based ECG signal quality assessment toolbox for a real-time QRS detector, followed by a spurious R-peak detector for signal processing and quality assurance. Then, the processed RR interval tachogram is fed to the above-described automated sleep staging algorithm. Sleep state classification can be performed with different levels of granularity, namely 2-level (Wake; Sleep), 3-level (Wake: NREM: N1+N2+N3; and REM) or 4-level (Wake; Light: N1+N2; Deep: N3; and REM) in 30 second epochs compliant with the American Academy of Sleep Medicine (AASM) standard. The 2 and 3 level classifications are obtained by appropriately collapsing the 4-level classification. Additional information describing the mode of operation of the algorithm are reported in the Supplement (see Supplement Figure 1).

**Figure 1.**
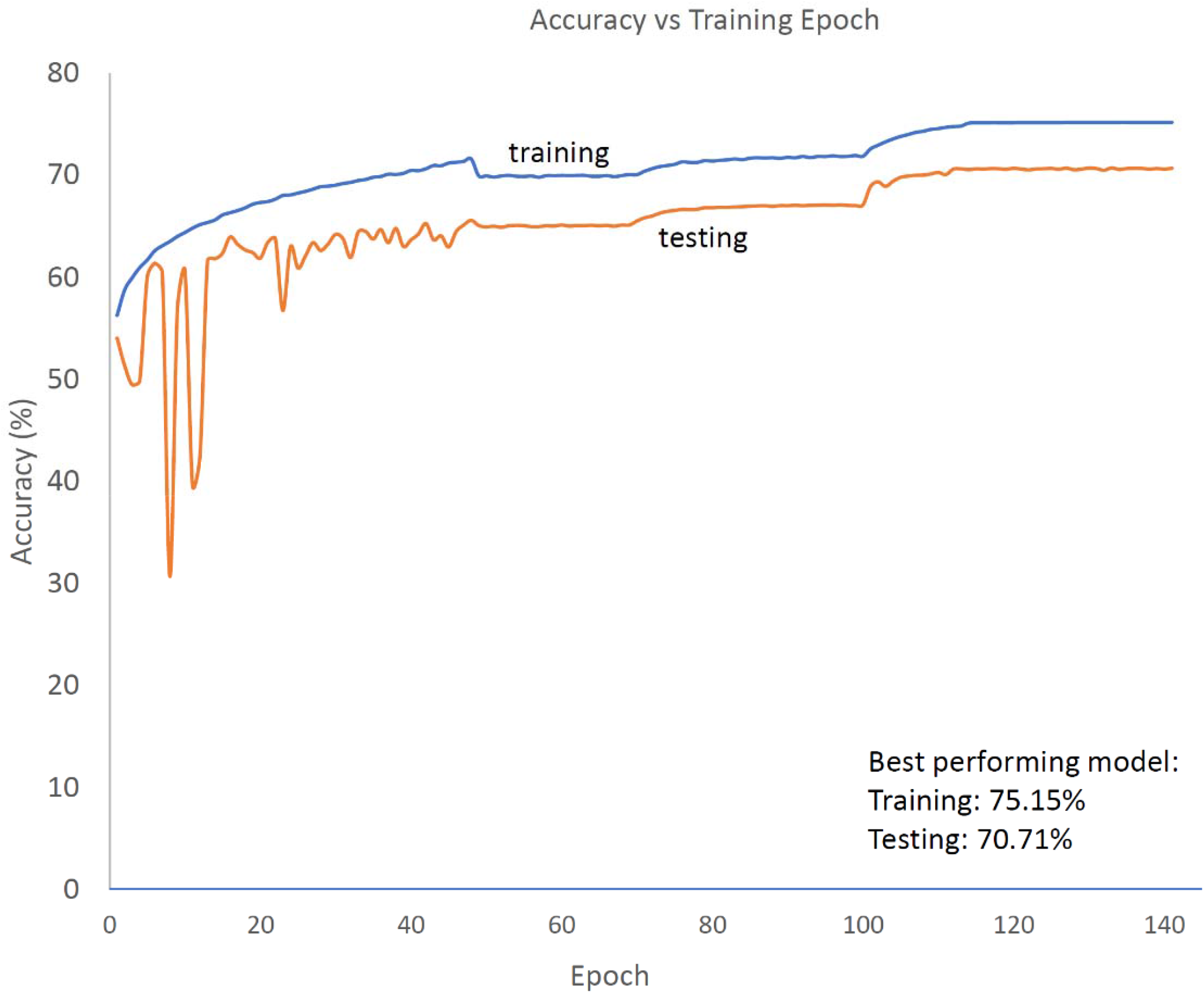
Trend of accuracy in the training (blue solid line) and testing (orange solid line) datasets as a function of training epochs for the 4-level model. Following an initialization period (for epoch value <∼50), the difference in performance between the training and testing sets stabilizes at ∼5%.

### Participants & Study Protocol

#### You Snooze You Win - The PhysioNet Computing in Cardiology (CinC) Challenge 2018 dataset

The CinC dataset is available at: https://physionet.org/content/challenge-2018/1.0.0/ and it is comprised of 994 participants (18-90 years old) which were monitored at Massachusetts General Hospital (MGH) sleep laboratory for the diagnosis of sleep disorders. Each participant has a complete set of 30-s annotated segments with corresponding sleep stages and respiratory events annotated by clinical staff at the MGH according to the AASM manual for the scoring of sleep. For this study, the initial preprocessing step consisted of assessing the quality of the ECG recordings extracted from the ensemble of recorded signals (EEG, EOG, EMG, respiration, and SaO_2_). The signal quality index is an estimate of signal to noise ratio (SNR). The ECG channel was fed to the Neurobit-HRV software to extract the RR interval tachogram along with a signal quality index. ECG traces with either a SNR <5 dB or with >10% unusable data were rejected and not included in the analytic steps described in the following sections.

#### Z3Pulse datasets

The Z3Pulse dataset consisted of a set of 52 healthy adults in the age range of 23 to 69 years with no known sleep or psychiatric disorders. All participants provided written informed consent in compliance with a protocol approved by the National University of Singapore’s Institutional Review Board (NUS-IRB) and were paid for their involvement. The study protocol was carried out over three nights, when trained research assistants visited participants’ homes. On each night, participants wore both a wearable ECG patch paired with an Android phone, alongside with simultaneous PSG. PSG was collected using the SOMNOtouch device (SOMNOmedics GmbH, Randersacker, Germany). Electroencephalography was recorded from two channels (C3 and C4 in the international 10–20 system of electrode placement) referenced to the contralateral mastoids. The common ground and reference electrode were placed at Fpz and Cz, respectively. Electrooculography (EOG; right and left outer canthi) and submental electromyography (EMG) were also recorded. EEG signals were sampled at 256 Hz and impedance was kept at less than 5KΩ for EEG and below 10KΩ for EOG and EMG channels. Sleep scoring was manually performed based on the AASM manual.^27^

Self-reported bed and wake up times were also collected. Each participant had at least one usable recording, 40 (∼77%) had 2 separate overnight recordings and 19 (∼37%) had 3.

The wearable ECG patch utilized in this study was the Z3Pulse device (Neurobit Inc., New York, USA), a chest worn, wireless device capable of recording HR, body position, activity and temperature. Z3Pulse was developed using the Movesense open wearable tech platform (Movesense, Finland, www.movesense.com). The device is first connected to a reusable belt worn around the chest or a one-time Ag-Cl patch (see Supplement Figure 2). In the latter case, the patch is directly applied on the skin below the sternum. The Movesense device consists of a single-channel ECG sensor, a nine-axis inertial measurement unit (IMU) and a temperature sensor. The device captures ECG at 128 Hz and IMU at 13 Hz. The Z3Pulse device transmits the data in real-time to a mobile app over Bluetooth low energy (BLE). Once recording is complete, the collected data is uploaded to the cloud and analyzed using Neurobit-HRV.

The process of aligning the data across PSG and Z3Pulse was essential to allow accurate comparison on an epoch-by-epoch basis. The timestamps of both systems were first synchronized using an internet time server as reference. Bedtimes for the PSG were estimated from the sleep diary. For Z3Pulse, position data were used to derive times on and off the bed. The time recorded from either the PSG or Z3Pulse, depending on the device with earlier lights-off time, was used as the reference data point. The shorter recording was extended to match the longer one by assuming “awake time” for missing epochs. An example of the scenario described is summarized in Figure S3 in the Supplement.

### Data Analysis

We first evaluated the classification performance of the 2-, 3-, and 4-levels models in both datasets considering each scored epoch as an independent observation. The goodness of fit metrics computed were the following: Accuracy (A), Cohen’s kappa (K), Sensitivity (SE), Specificity (SP), Positive Predictive Value (PPV), and Negative Predicted Value (NPV). Then, the estimates obtained at the epoch level were averaged for each participant, to derive individual metrics and the same goodness of fit metrics were computed.

The wealth of data within the CinC dataset allowed for performance evaluation of the algorithm as a function of factors known to have an impact on HR; age, AHI score, and gender. Participants’ ages and AHI scores were independently stratified using 3-level categorization; age≤40 years-old, 40<age≤60 years-old, or age>60 years-old and AHI≤5 (None/Minimal) or 5<AHI≤15 (Mild) or AHI>15 (Severe). Multiple independent linear regression models were used to estimate the association between age, AHI score, and gender on A, K, SE, SP, PPV, and NPV. The limited sample size of the Z3Pulse dataset did not allow for analyses stratified by age, AHI score, and gender.

## RESULTS

### Populations

For the CinC dataset a total of 6 participants were excluded from the analyses as the associated ECG traces had either a SNR <5 dB or >10% unusable data, thus the final dataset included 988 subjects. There were 664 (∼67%) males, 324 (∼33%) females; 137 (∼14%), 285 (∼29%) and 566 (∼57%) participants in the None/Minimal, Mild, and Severe AHI groups, respectively; 157 (∼16%), 456 (∼46%) and 375 (∼38%) participants in the age≤40 years-old, 40<age≤60 years-old, and age>60 years-old groups, respectively.

For the Z3pulse dataset, there were 27 (∼52%) males, 25 (∼48%) females; 38 (∼73%), 7 (∼13%) and 7 (∼13%) participants in the age≤40 years-old, 40<age≤60 years-old, and age>60 years-old groups, respectively.

### CinC dataset validation

Results for the 2-, 3-, and 4-levels prediction for the CinC dataset data are displayed in Figure 2 via confusion matrices and reported in **Table SA**. The highest value of accuracy was achieved by the 2-levels classification (wake vs sleep), whereas the predictions obtained in the 3-levels model obtained the best value of K. By design of the algorithm, all the derived metrics (SE, SP, PPV, and NPV) relative to the classification of wake segments, are identical across the 2-, 3-, and 4-levels models. On the other hand, the classification of sleep segments is achieved by progressively collapsing different sleep states passing from the 4-levels to the 2-levels model. As a consequence, values of PPV and NPV were substantially equivalent across models and sleep states. The granular level of classification achieved by the 4-levels model obtained the lowest value of SE (0.3812) and the highest value of SP (0.9744) for the classification of DEEP sleep segments. The comparison of the goodness of fit metrics at the segment level (**Table SA**) versus the ones obtained at the participant level, (**Table A**) revealed a close concordance between the two approaches. In addition, the narrow confidence intervals (ranging from ±1% to ±5% of the mean) of the averaged metrics in **Table A** indicates the absence of underfitting nor overfitting.

**Table A.**
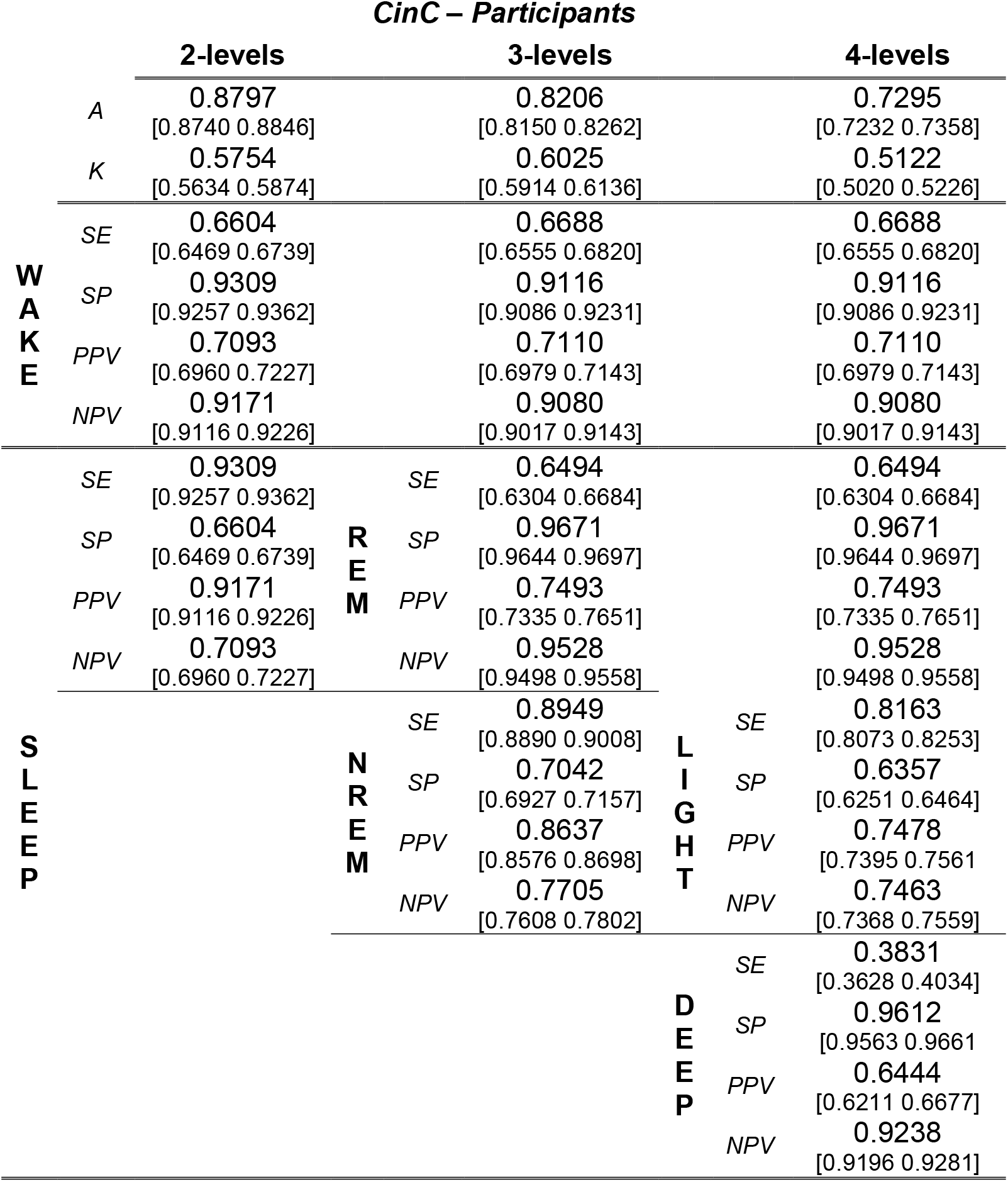
Mean and [95% Confidence Intervals] Accuracy (A), Cohen’s kappa (K), Sensitivity (SE), Specificity (SP), Positive Predictive Value (PPV), and Negative Predicted Value (NPV) were obtained by firstly collapsing metrics within and subsequently across participants in the CinC dataset. Results are reported separately for the 2-, 3- and 4-levels models.

**Figure 2.**
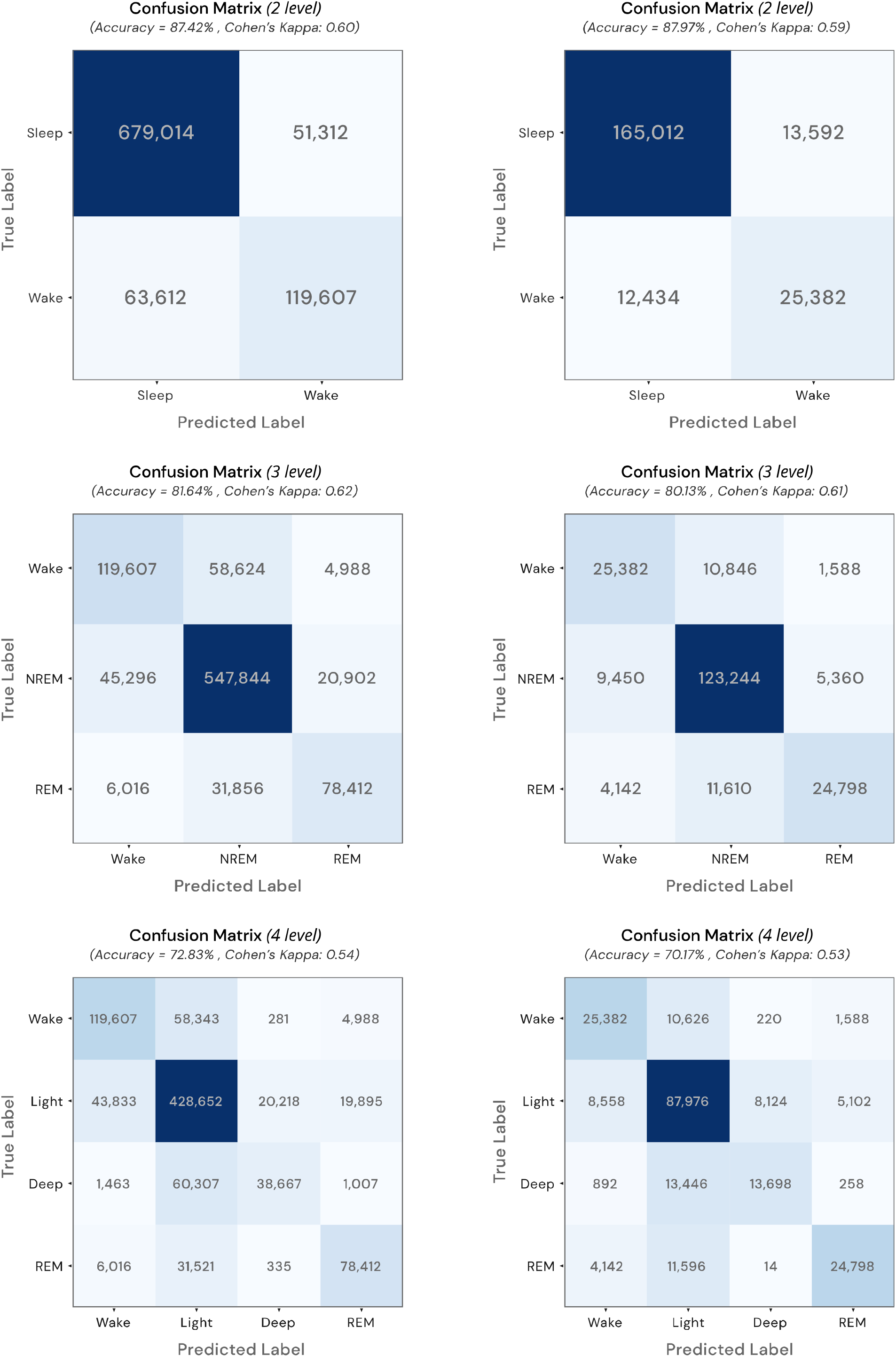
Confusion matrices for results for the 2-, 3-, and 4-levels prediction for the CinC dataset

**Table B** summarizes the performance of the 2-levels model as a function of participants’ characteristics. AHI and gender groups did not affect the outcome metrics. In contrast, a significant decrease in A and K were reported for older age groups when compared to the reference age group (age≤40 years-old). Specifically, the average decrease in A for the age group 40<Age≤60 and Age>60 was 3.02% and 7.45%, respectively. Similar results were found for the other outcome metrics such that the mean decrease in performance in the group of participants age>60 years old is approximately double that of the 40<age≤60 group. Analogous findings were reported when considering the 3-levels and 4-levels models as reported in **Table SB** and **Table SC**.

**Table B.**
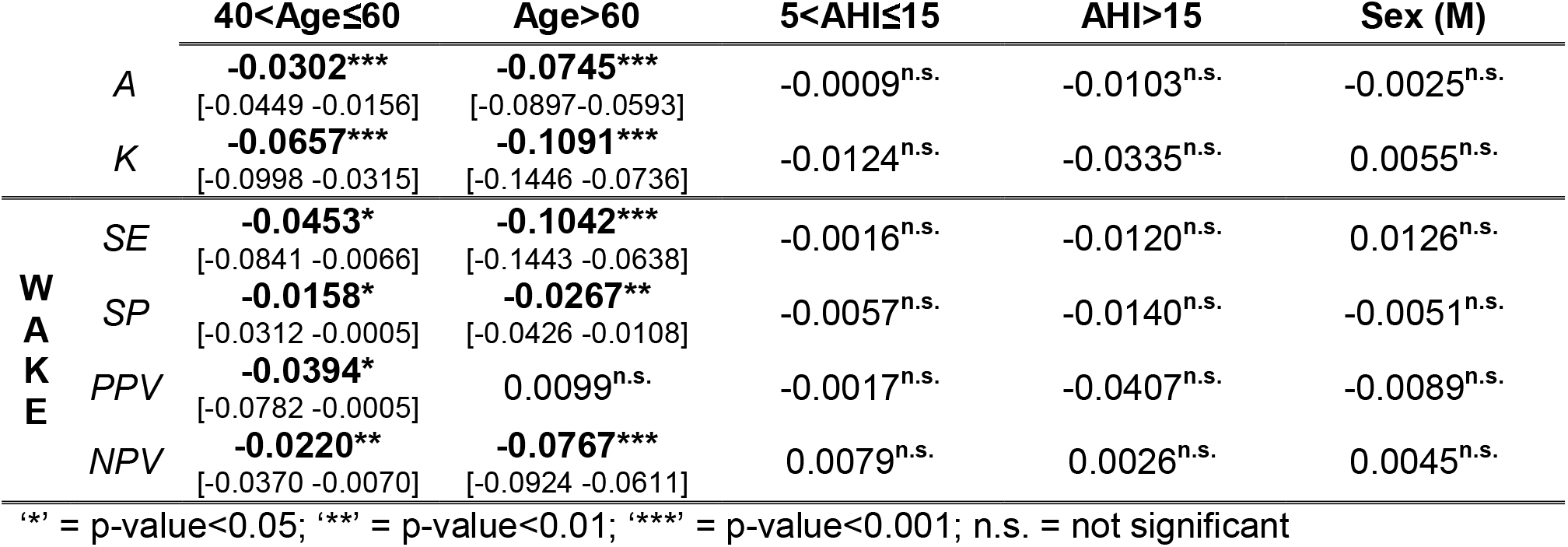
Mean and [95% Confidence Intervals] of beta estimates of the performance of the 2-levels models as a function of the participants’ characteristics: age, AHI score, and biological sex.

### Z3Pulse dataset

Results for the 2-, 3-, and 4-levels prediction are reported in **Table C** (at the participant level) and **Table SD** (at the segment level). The highest value of accuracy was achieved by the binary classification (wake vs sleep) performed in the 2-levels model (0.8812 [0.8668-0.8955] and 0.8797, at the participant and segment levels, respectively), whereas the prediction obtained in the 3-levels model obtained the best value of K (0.611 [0.5818 0.6401] and 0.6117]. Regarding the classification of the sleep states, the lowest value of SE (0.6163 [0.5710 0.6617] and 0.6115) and the highest value of SP (0.9606 [0.9536 0.9676] and 0.9605) were obtained across the classification of DEEP sleep segment. Values of PPV and NPV were comparable equivalent across models and sleep states. The comparison of the goodness of fit metrics at the segment (**Table C**) versus that obtained at the participant level (**Table SD**) revealed a close concordance across the different metrics.

**Table C.**
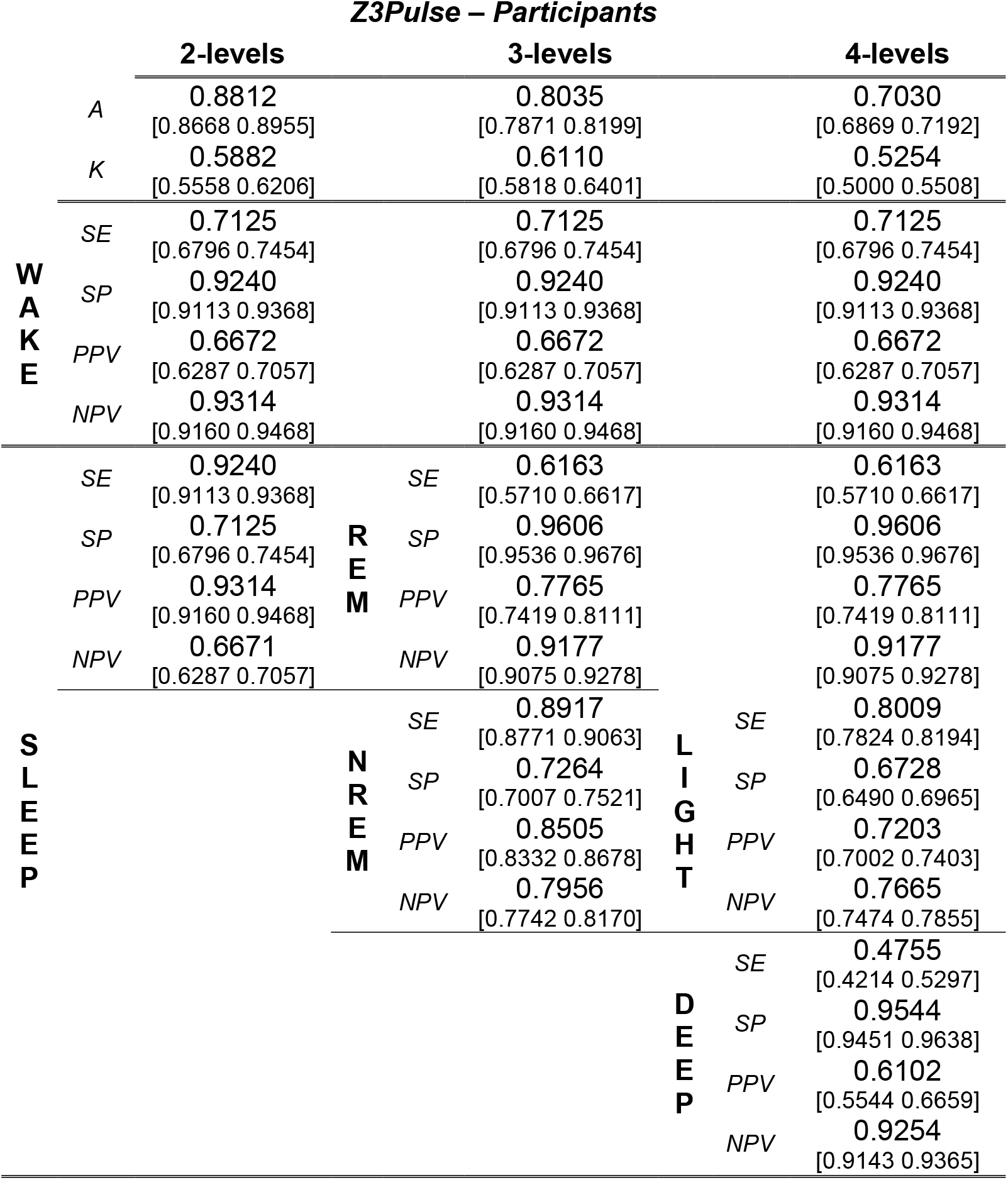
Mean and [95% Confidence Intervals] Accuracy (A), Cohen’s kappa (K), Sensitivity (SE), Specificity (SP), Positive Predictive Value (PPV), and Negative Predicted Value (NPV) were obtained by firstly collapsing metrics within and subsequently across participants in the Z3Pulse dataset. Results are reported separately for the 2-, 3- and 4-levels models.

## DISCUSSION

The aim of this study was to validate the performance of the Neurobit-HRV Deep Learning models for automatic sleep staging, utilizing two datasets, namely a publicly available collection of PSG recordings (You Snooze You Win - The PhysioNet CinC Challenge 2018 dataset^25^), and a proprietary dataset of simultaneously collected PSG and single lead ECG acquired using a wearable device (Z3Pulse device). The results of the classification performance in the two datasets highlighted high and consistent accuracy, 88%, 82% and 73% for the 2 levels, 3 levels and 4 levels models respectively in the CinC dataset and 88%, 80% and 70% for the Z3Pulse dataset. In addition, the agreement of such metrics compared to results obtained in the training/testing phase (Figure 1), speaks to the robustness of the developed framework. In fact, the accuracy of the 4-level model in the original training (75.15%) and testing set (70.71%) is comparable to the results obtained in the CinC (72.95%) and Z3Pulse (70.30%) datasets.

Recent years have seen the surge of automated sleep stage algorithms leveraging a variety of physiological signals collected via traditional, semiportable, or wearables devices. While models trained on the entirety of the physiological signals collected via PSG are able to achieve performance close to human scorers,^28^ the interest in quantifying and tracking human sleep by means of less invasive and easy deployable devices has increased exponentially. Therefore, the demand for automated, accurate, and highspeed algorithms working with a reduced set of features has scaled accordingly. Proposed models have evolved from simple binary detection of sleep–wake to a more fine-grained sleep classification. Examples are automatic frameworks utilizing a single lead EEG, either isolated from PSG recordings or collected using the Zmachine^®^ sleep monitoring system,^29–31^ or a single lead ECG either isolated from PSG studies or acquired using a variety of strategies^12,32,33^ potentially complemented by respiratory effort ^34,35^ The model validated in this study provides a significant contribution toward the methodological advancement of automatic sleep stage scoring relying solely on a single lead ECG signal, acquired via a wearable patch. In addition, to fully describe the performance of the algorithm, we calculated several goodness of fit in addition to accuracy, such as K, SP, SE, NPV and PPV not only for the overall models but for each of the predicted sleep states. Although these values are inextricably linked, they do provide complementary information^36^ and allow for a better assessment of the performance of the algorithm.

One advantage of the proposed algorithm is that it does not require contextual information, such as time of the recording, thus being agnostic and removing any potential sources of bias. This type of bias is known to affect algorithms to analyze actigraphy data^37^ and manual PSG scoring. We also tested if other demographic factors known to be associated with sleep characteristics, namely gender, age and AHI, would affect algorithm performances. We showed the insensitivity of the algorithm to both AHI and gender. Such characteristics have been previously reported to affect classification performance^38^ thus, our methodology is seen as an advancement toward a universally applicable sleep scoring model. However, our results showed that the performance of the algorithm was associated with participants’ age. One explanation for this finding may be attributed to the differences in age group distributions between the training/testing versus the validation sets. Extensive literature has shown that HR indices parameters change with age,^39^ thus for future development we anticipate additional data collection across various age groups to optimize the algorithm.

Lastly, the algorithm and datasets utilized in this paper are publicly available and we believe this is an essential step to support wide adoption of automatic sleep staging algorithms within the research and clinical communities.

In conclusion, result presented in this paper highlight the possibility of utilizing inexpensive and widely available ECG wearable deceives paired with automated sleep staging algorithms to characterize sleep architecture easily and inexpensively with high levels of accuracy. This would open the possibility of new scalable and accessible modalities for monitoring sleep in the home environment, increasing equitable access to sleep health.

## Data Availability

The dataset You Snooze You Win - The PhysioNet Computing in Cardiology Challenge 2018 is available online at https://physionet.org/content/challenge-2018/1.0.0/.
The dataset Z3Pulse is available upon reasonable request to the authors, subject to review by an ethics committee.

## Acknowledgements

This work was supported by grants from the Health Technology Consortium grant. The authors would also like to thank Xinyu Chua, Andrew Dicom, Shohreh Ghorbani, Hosein Golkashani, Zuriel Hassirim, Shamsul Azrin Bin Jamaludin, Tiffany Koa, Teyang Lau, Zaven Leow, Karthika Muthiah, Alyssa Ng, Raelynn Ng, Zhenghao Pu, Teck Boon Teo, Kian Wong and Nicole Yu for their help in data collection and sleep scoring.

## Preprint

The manuscript is currently deposited on Medrxiv.

## REFERENCES

1. Gottlieb DJ, Redline S, Nieto FJ, et al. Association of usual sleep duration with hypertension: the Sleep Heart Health Study. Sleep. 2006;29(8):1009–1014. doi:10.1093/sleep/29.8.1009

2. Grandner MA, Alfonso-Miller P, Fernandez-Mendoza J, Shetty S, Shenoy S, Combs D. Sleep: important considerations for the prevention of cardiovascular disease. Curr Opin Cardiol. 2016;31(5):551.

3. St-Onge M. Sleep–obesity relation: underlying mechanisms and consequences for treatment. Obes Rev. 2017;18:34–39.

4. Alvaro PK, Roberts RM, Harris JK. A systematic review assessing bidirectionality between sleep disturbances, anxiety, and depression. Sleep. 2013;36(7):1059–1068.

5. Malhotra RK. Neurodegenerative disorders and sleep. Sleep Med Clin. 2018;13(1):63–70.

6. Altevogt BM, Colten HR. Sleep Disorders and Sleep Deprivation: An Unmet Public Health Problem. National Academies Press; 2006.

7. Ohayon MM. Epidemiological overview of sleep disorders in the general population. Sleep Med Res. 2011;2(1):1–9.

8. Russo K, Goparaju B, Bianchi MT. Consumer sleep monitors: is there a baby in the bathwater? Nat Sci Sleep. 2015;7:147–157. doi:10.2147/NSS.S94182

9. Fuller D, Colwell E, Low J, et al. Reliability and validity of commercially available wearable devices for measuring steps, energy expenditure, and heart rate: systematic review. JMIR mHealth uHealth. 2020;8(9):e18694.

10. de Zambotti M, Cellini N, Menghini L, Sarlo M, Baker FC. Sensors Capabilities, Performance, and Use of Consumer Sleep Technology. Sleep Med Clin. 2020;15(1):1–30. doi:10.1016/j.jsmc.2019.11.003

11. Menghini L, Yuksel D, Goldstone A, Baker FC, de Zambotti M. Performance of Fitbit Charge 3 against polysomnography in measuring sleep in adolescent boys and girls. Chronobiol Int. 2021;38(7):1010–1022. doi:10.1080/07420528.2021.1903481

12. Sridhar N, Shoeb A, Stephens P, et al. Deep learning for automated sleep staging using instantaneous heart rate. NPJ Digit Med. 2020;3(1):1–10.

13. Fonseca P, Weysen T, Goelema MS, et al. Validation of photoplethysmography-based sleep staging compared with polysomnography in healthy middle-aged adults. Sleep. 2017;40(7).

14. Li Q, Li Q, Cakmak AS, et al. Transfer learning from ECG to PPG for improved sleep staging from wrist-worn wearables. Physiol Meas. 2021;42(4):44004.

15. de Zambotti M, Baker FC. Sleep and Circadian Regulation of the Autonomic Nervous System. Auton Nerv Syst Sleep. 2021;5:63.

16. Aldredge JL, Welch AJ. Variations of heart rate during sleep as a function of the sleep cycle. Electroencephalogr Clin Neurophysiol. 1973;35(2):193–198.

17. Mendez M, Bianchi AM, Villantieri O, Cerutti S. Time-varying analysis of the heart rate variability during REM and non REM sleep stages. In: 2006 International Conference of the IEEE Engineering in Medicine and Biology Society. IEEE; 2006:3576–3579.

18. Versace F, Mozzato M, Tona GDM, Cavallero C, Stegagno L. Heart rate variability during sleep as a function of the sleep cycle. Biol Psychol. 2003;63(2):149–162.

19. Radha M, Fonseca P, Moreau A, et al. A deep transfer learning approach for wearable sleep stage classification with photoplethysmography. npj Digit Med. 2021;4(1):135. doi:10.1038/s41746-021-00510-8

20. Wulterkens BM, Fonseca P, Hermans LWA, et al. It is all in the wrist: wearable sleep staging in a clinical population versus reference polysomnography. Nat Sci Sleep. 2021;13:885.

21. Lu G, Yang F. Limitations of Oximetry to Measure Heart Rate Variability Measures. Cardiovasc Eng. 2009;9(3):119–125. doi:10.1007/s10558-009-9082-3

22. Jan H-Y, Chen M-F, Fu T-C, Lin W-C, Tsai C-L, Lin K-P. Evaluation of Coherence Between ECG and PPG Derived Parameters on Heart Rate Variability and Respiration in Healthy Volunteers With/Without Controlled Breathing. J Med Biol Eng. 2019;39(5):783–795. doi:10.1007/s40846-019-00468-9

23. Lee XK, Chee Niyn, Ong JL, et al. Validation of a consumer sleep wearable device with actigraphy and polysomnography in adolescents across sleep opportunity manipulations. J Clin Sleep Med. 2019;15(9):1337–1346.

24. Depner CM, Cheng PC, Devine JK, et al. Wearable technologies for developing sleep and circadian biomarkers: a summary of workshop discussions. Sleep. 2020;43(2):zsz254.

25. Goldberger AL, Amaral LAN, Glass L, et al. PhysioBank, PhysioToolkit, and PhysioNet: components of a new research resource for complex physiologic signals. Circulation. 2000;101(23):e215–e220.

26. https://gist.github.com/neurobittechnologies/e1d13e0b59c56f5804af7c127b36d20d.

27. Silber MH, Ancoli-Israel S, Bonnet MH, et al. The visual scoring of sleep in adults. J Clin sleep Med. 2007;3(02):121–131.

28. Vallat R, Walker MP. An open-source, high-performance tool for automated sleep staging. Elife. 2021;10:e70092.

29. Wang Y, Loparo KA, Kelly MR, Kaplan RF. Evaluation of an automated single-channel sleep staging algorithm. Nat Sci Sleep. 2015;7:101.

30. Yoon H, Hwang SH, Choi J-W, Lee YJ, Jeong D-U, Park KS. REM sleep estimation based on autonomic dynamics using R–R intervals. Physiol Meas. 2017;38(4):631.

31. Olesen AN, Jørgen Jennum P, Mignot E, Sorensen HBD. Automatic sleep stage classification with deep residual networks in a mixed-cohort setting. Sleep. 2021;44(1):zsaa161. doi:10.1093/sleep/zsaa161

32. Li Q, Li Q, Liu C, Shashikumar SP, Nemati S, Clifford GD. Deep learning in the cross-time frequency domain for sleep staging from a single-lead electrocardiogram. Physiol Meas. 2018;39(12):124005.

33. Walch O, Huang Y, Forger D, Goldstein C. Sleep stage prediction with raw acceleration and photoplethysmography heart rate data derived from a consumer wearable device. Sleep. 2019;42(12):zsz180.

34. Fonseca P, Long X, Radha M, Haakma R, Aarts RM, Rolink J. Sleep stage classification with ECG and respiratory effort. Physiol Meas. 2015;36(10):2027.

35. Bakker JP, Ross M, Vasko R, et al. Estimating sleep stages using cardiorespiratory signals: validation of a novel algorithm across a wide range of sleep-disordered breathing severity. J Clin Sleep Med. 2021:jcsm-9192.

36. Trevethan R. Sensitivity, Specificity, and Predictive Values: Foundations, Pliabilities, and Pitfalls in Research and Practice. Front Public Heal. 2017;5:307.

37. Paquet J, Kawinska A, Carrier J. Wake detection capacity of actigraphy during sleep. Sleep. 2007;30(10):1362–1369.

38. Redline S, Kirchner HL, Quan SF, Gottlieb DJ, Kapur V, Newman A. The Effects of Age, Sex, Ethnicity, and Sleep-Disordered Breathing on Sleep Architecture. Arch Intern Med. 2004;164(4):406–418. doi:10.1001/archinte.164.4.406

39. Voss A, Schroeder R, Heitmann A, Peters A, Perz S. Short-term heart rate variability—influence of gender and age in healthy subjects. PLoS One. 2015;10(3):e0118308.

