## Supplement for "An automated heart rate-based algorithm for sleep stage classification: validation using conventional PSG and innovative wearable ECG device"

**SUPPLEMENTARY TABLES**

**Table SA** – Mean Accuracy (A), Cohen's kappa (K), Sensitivity (SE), Specificity (SP), Positive Predictive Value (PPV), and Negative Predicted Value (NPV) averaged across the available segments in the CinC dataset. Results are reported separately for the 2-, 3- and 4-levels models.

|  | ***CinC –Segments*** | | | | | | | |
| --- | --- | --- | --- | --- | --- | --- | --- | --- |
|  |  | **2-levels** |  |  | **3-levels** |  |  | **4-levels** |
|  | *A* | 0.8742 |  |  | 0.8146 |  |  | 0.7283 |
|  | *K* | 0.5976 |  |  | 0.6162 |  |  | 0.5357 |
| **W**  **A**  **K**  **E** | *SE* | 0.6528 |  |  | 0.6528 |  |  | 0.6528 |
|  | *SP* | 0.9297 |  |  | 0.9297 |  |  | 0.9297 |
|  | *PPV* | 0.6998 |  |  | 0.6998 |  |  | 0.6998 |
|  | *NPV* | 0.9143 |  |  | 0.9143 |  |  | 0.9143 |
| **S**  **L**  **E**  **E**  **P** | *SE* | 0.9297 | **R**  **E**  **M** | *SE* | 0.6743 |  |  | 0.6743 |
|  | *SP* | 0.6528 |  | *SP* | 0.9675 |  |  | 0.9675 |
|  | *PPV* | 0.9143 |  | *PPV* | 0.7518 |  |  | 0.7518 |
|  | *NPV* | 0.6998 |  | *NPV* | 0.9532 |  |  | 0.9532 |
|  |  |  | **N**  **R**  **E**  **M** | *SE* | 0.8922 | **L**  **I**  **G**  **H**  **T** | *SE* | 0.8362 |
|  |  |  |  | *SP* | 0.6979 |  | *SP* | 0.6255 |
|  |  |  |  | *PPV* | 0.8583 |  | *PPV* | 0.7406 |
|  |  |  |  | *NPV* | 0.7595 |  | *NPV* | 0.7492 |
|  |  |  |  |  |  | **D**  **E**  **E**  **P** | *SE* | 0.3812 |
|  |  |  |  |  |  |  | *SP* | 0.9744 |
|  |  |  |  |  |  |  | *PPV* | 0.6499 |
|  |  |  |  |  |  |  | *NPV* | 0.9265 |

**Table SB** – Mean and [95% Confidence Intervals] of beta estimates of the performance of the 3-levels models as a function of the participants’ characteristics: age, AHI score, and gender.

|  |  | **40<Age≤60** | **Age>60** | **5<AHI≤15** | **AHI>15** | **Gender (M)** |
| --- | --- | --- | --- | --- | --- | --- |
|  | *A* | **-0.0287*****  [-0.0443 -0.0131] | **-0.0735*****  [-0.0896 -0.0573] | -0.0106**^n.s.^** | -0.0144**^n.s.^** | -0.0083**^n.s.^** |
|  | *K* | **-0.0403****  [-0.0710 -0.0096] | **-0.1348*****  [-0.1666 -0.1029] | -0.0228**^n.s.^** | -0.0238**^n.s.^** | 0.0063**^n.s.^** |
| **W**  **A**  **K**  **E** | *SE* | **-0.0405***  [-0.07890 -0.0020] | **-0.0956*****  [-0.1354 -0.0559] | -0.0019**^n.s.^** | -0.0008**^n.s.^** | 0.0112**^n.s.^** |
|  | *SP* | **-0.0291*****  [-0.0501 -0.0082] | **-0.0461****  [-0.0677 -0.0244] | -0.0005**^n.s.^** | -0.0169**^n.s.^** | -0.0151**^n.s.^** |
|  | *PPV* | **-0.0434***  [-0.0818 -0.0051] | 0.0014**^n.s.^** | -0.0109**^n.s.^** | -0.0298**^n.s.^** | -0.0092**^n.s.^** |
|  | *NPV* | **-0.0188****  [-0.0365 -0.0010] | **-0.0791*****  [-0.0973 -0.0608] | 0.0100**^n.s.^** | 0.0007**^n.s.^** | 0.0006**^n.s.^** |
| **R**  **E M** | *SE* | -0.0188**^n.s.^** | **-0.2026*****  [-0.2576 -0.1478] | -0.0070**^n.s.^** | -0.0026**^n.s.^** | 0.0484**^n.s.^** |
|  | *SP* | -0.0028**^n.s.^** | 0.0066**^n.s.^** | -0.0097**^n.s.^** | -0.0061**^n.s.^** | **-0.0109*****  [-0.0165 -0.0052] |
|  | *PPV* | -0.0038**^n.s.^** | **-0.0700*****  [-0.1172 -0.0228] | -0.0245**^n.s.^** | -0.0130**^n.s.^** | -0.0403**^n.s.^** |
|  | *NPV* | -0.0005**^n.s.^** | **-0.0180***  [-0.0199 -0.0017] | -0.0034**^n.s.^** | -0.0306**^n.s.^** | -0.0007**^n.s.^** |
| **N**  **R**  **E M** | *SE* | **-0.0203***  [-0.0374 -0.0032] | -0.0146**^n.s.^** | -0.0142**^n.s.^** | -0.0188**^n.s.^** | 0.0181**^n.s.^** |
|  | *SP* | -0.0196**^n.s.^** | **-0.1320*****  [-0.1650 -0.0989] | -0.0002**^n.s.^** | -0.0048**^n.s.^** | -0.0307**^n.s.^** |
|  | *PPV* | **-0.0225*****  [-0.0391 -0.0059] | **-0.0872*****  [-0.1044 -0.0699] | 0.0057**^n.s.^** | 0.0021**^n.s.^** | 0.0060**^n.s.^** |
|  | *NPV* | -0.0149**^n.s.^** | 0.0034**^n.s.^** | -0.0285**^n.s.^** | -0.0332**^n.s.^** | -0.0144**^n.s.^** |

‘*’ = p-value<0.05; ‘**’ = p-value<0.01; ‘***’ = p-value<0.001; n.s. = not significant

**Table SC** – Mean and [95% Confidence Intervals] of beta estimates of the performance of 4-levels models as a function of the participants’ characteristics: age, AHI score, and gender.

|  |  | **40<Age≤60** | **Age>60** | **5<AHI≤15** | **AHI>15** | **Gender (M)** |
| --- | --- | --- | --- | --- | --- | --- |
|  | *A* | **-0.0287*****  [-0.0443 -0.0131] | **-0.0735*****  [-0.0896 -0.0573] | -0.0106**^n.s.^** | -0.0144**^n.s.^** | -0.0083**^n.s.^** |
|  | *K* | **-0.0403****  [-0.0710 -0.0096] | **-0.1348*****  [-0.1666 -0.1029] | -0.0228**^n.s.^** | -0.0238**^n.s.^** | 0.0063**^n.s.^** |
| **W**  **A**  **K**  **E** | *SE* | **-0.0405***  [-0.07890 -0.0020] | **-0.0956*****  [-0.1354 -0.0559] | -0.0019**^n.s.^** | -0.0008**^n.s.^** | 0.0112**^n.s.^** |
|  | *SP* | **-0.0291*****  [-0.0501 -0.0082] | **-0.0461****  [-0.0677 -0.0244] | -0.0005**^n.s.^** | -0.0169**^n.s.^** | -0.0151**^n.s.^** |
|  | *PPV* | **-0.0434***  [-0.0818 -0.0051] | 0.0014**^n.s.^** | -0.0109**^n.s.^** | -0.0298**^n.s.^** | -0.0092**^n.s.^** |
|  | *NPV* | **-0.0188****  [-0.0365 -0.0010] | **-0.0791*****  [-0.0973 -0.0608] | 0.0100**^n.s.^** | 0.0007**^n.s.^** | 0.0006**^n.s.^** |
| **R**  **E M** | *SE* | -0.0188**^n.s.^** | **-0.2026*****  [-0.2576 -0.1478] | -0.0070**^n.s.^** | -0.0026**^n.s.^** | 0.0484**^n.s.^** |
|  | *SP* | -0.0028**^n.s.^** | 0.0066**^n.s.^** | -0.0097**^n.s.^** | -0.0061**^n.s.^** | **-0.0109*****  [-0.0165 -0.0052] |
|  | *PPV* | -0.0038**^n.s.^** | **-0.0700*****  [-0.1172 -0.0228] | -0.0245**^n.s.^** | -0.0130**^n.s.^** | -0.0403**^n.s.^** |
|  | *NPV* | -0.0005**^n.s.^** | **-0.0180***  [-0.0199 -0.0017] | -0.0034**^n.s.^** | -0.0306**^n.s.^** | -0.0007**^n.s.^** |
| **L**  **I**  **G**  **H**  **T** | *SE* | -0.0235**^n.s.^** | -0.0179**^n.s.^** | 0.0053**^n.s.^** | 0.0077**^n.s.^** | -0.0131**^n.s.^** |
|  | *SP* | -0.0225**^n.s.^** | **-0.1180*****  [-0.1490 -0.0871] | 0.0072**^n.s.^** | 0.0076**^n.s.^** | **0.0337*****  [0.0118 0.0557] |
|  | *PPV* | -0.0225**^n.s.^** | **-0.0699*****  [-0.0943 -0.0455] | 0.0103**^n.s.^** | 0.0248**^n.s.^** | **0.0436*****  [0.0269 0.0610] |
|  | *NPV* | -0.0270**^n.s.^** | -0.0214**^n.s.^** | 0.0013**^n.s.^** | 0.0043**^n.s.^** | -0.0252**^n.s.^** |
| **D E**  **E**  **P** | *SE* | **-0.0971*****  [-0.1548 -0.0394] | **-0.2144*****  [-0.2741 -0.1548] | 0.0334**^n.s.^** | -0.0298**^n.s.^** | -0.0141**^n.s.^** |
|  | *SP* | 0.0045**^n.s.^** | 0.0044**^n.s.^** | 0.0114**^n.s.^** | 0.0133**^n.s.^** | 0.0017**^n.s.^** |
|  | *PPV* | **-0.0657***  [-0.1301 -0.0013] | **-0.1197*****  [-0.1885 -0.0508] | 0.0445**^n.s.^** | 0.0463**^n.s.^** | -0.0519**^n.s.^** |
|  | *NPV* | -0.0022**^n.s.^** | 0.0025**^n.s.^** | -0.0010**^n.s.^** | 0.0035**^n.s.^** | **0.0245*****  [0.0154 0.0336] |

‘*’ = p-value<0.05; ‘**’ = p-value<0.01; ‘***’ = p-value<0.001; n.s. = not significant

**Table SD** – Mean Accuracy (A), Cohen's kappa (K), Sensitivity (SE), Specificity (SP), Positive Predictive Value (PPV), and Negative Predicted Value (NPV) averaged across the available segments in the Z3Pulse dataset. Results are reported separately for the 2-, 3- and 4-levels models.

|  | ***Z3Pulse – Segments*** | | | | | | | |
| --- | --- | --- | --- | --- | --- | --- | --- | --- |
|  |  | **2-levels** |  |  | **3-levels** |  |  | **4-levels** |
|  | *A* | 0.8797 |  |  | 0.8013 |  |  | 0.7017 |
|  | *K* | 0.5880 |  |  | 0.6117 |  |  | 0.5324 |
| **W**  **A**  **K**  **E** | *SE* | 0.6712 |  |  | 0.6712 |  |  | 0.6712 |
|  | *SP* | 0.9239 |  |  | 0.9239 |  |  | 0.9239 |
|  | *PPV* | 0.6513 |  |  | 0.6513 |  |  | 0.6513 |
|  | *NPV* | 0.9299 |  |  | 0.9299 |  |  | 0.9299 |
| **S**  **L**  **E**  **E**  **P** | *SE* | 0.9239 | **R**  **E**  **M** | *SE* | 0.6115 |  |  | 0.6115 |
|  | *SP* | 0.6712 |  | *SP* | 0.9605 |  |  | 0.9605 |
|  | *PPV* | 0.9299 |  | *PPV* | 0.7811 |  |  | 0.7811 |
|  | *NPV* | 0.6513 |  | *NPV* | 0.9147 |  |  | 0.9147 |
|  |  |  | **N**  **R**  **E**  **M** | *SE* | 0.8927 | **L**  **I**  **G**  **H**  **T** | *SE* | 0.8015 |
|  |  |  |  | *SP* | 0.7134 |  | *SP* | 0.6656 |
|  |  |  |  | *PPV* | 0.8459 |  | *PPV* | 0.7115 |
|  |  |  |  | *NPV* | 0.7906 |  | *NPV* | 0.9249 |
|  |  |  |  |  |  | **D**  **E**  **E**  **P** | *SE* | 0.4841 |
|  |  |  |  |  |  |  | *SP* | 0.9556 |
|  |  |  |  |  |  |  | *PPV* | 0.6211 |
|  |  |  |  |  |  |  | *NPV* | 0.9249 |

**SUPPLEMENTARY FIGURES**

**Figure S1** - The algorithm operates on windows of length equal to 36 minutes (equivalent to 72 epochs of duration 30 seconds). In the scenario of a recording of this duration, the scores (on an epoch-by-epoch base) are obtained for 64 epochs (epoch 5 through 68) excluding the initial and final 4 epochs, resulting in a total of 8 epochs (4 minutes) not scored. The sleep state of the unscored epochs is assumed to be the same of the first and last scores segment, respectively. When the duration of a given recording exceeds 36 minutes, the 72-epoch window is progressively slid (in blocks of 64 epochs) to score the entirety of such recording. Lastly, if the number of epochs in a given recording are not multiple of 64, the window is slid to a quantity to allow complete coverage of the data.

A schematic view of the illustrated process is displayed below.

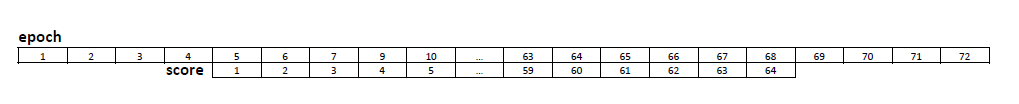

**Figure S2** Z3Pulse device

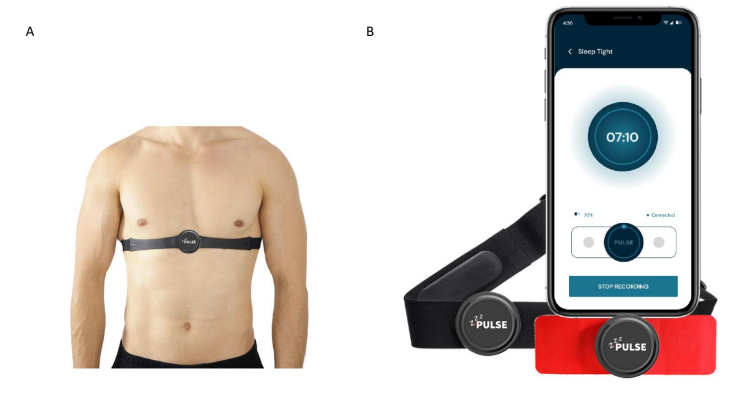

**Figure S3** – By study design, recordings from both devices are allowed to start at different times (within a reasonable offset). To carry out the proposed epoch-by-epoch comparison, it is required to align the measurements derived from the two systems. A scenario is presented in the following. In this example, the *lights off time* recorded by the PSG system was 21:00:10 [hh:mm:ss] (09:00:10PM) whereas the Z3Pulse *lights off time* was at 21:01:30 (09:01:30PM). The next mornig, the Z3Pulse recording ended three epochs earlier at 07:02:00 (07:02:00AM) the PSG recording, 07:03:40 (07:30:40AM). In this scenario, the PSG system is assumed as the reference given that the data recording with this system starts earlier than the Z3Pulse. While the two recordings are not aligned at the second level, nearest match is obtained by aligning the 1^st^ epoch of Z3Pulse recording with the 4^th^ one of the PSG signal. Once alignment is finalized, the duration of the two overnight recordings is normalized by assuming wake for missing epochs (either at the beginning or at the end of the recording(s)). In the discussed scenario, the first and last three epochs for Z3Pulse systems are assumed to be wake.

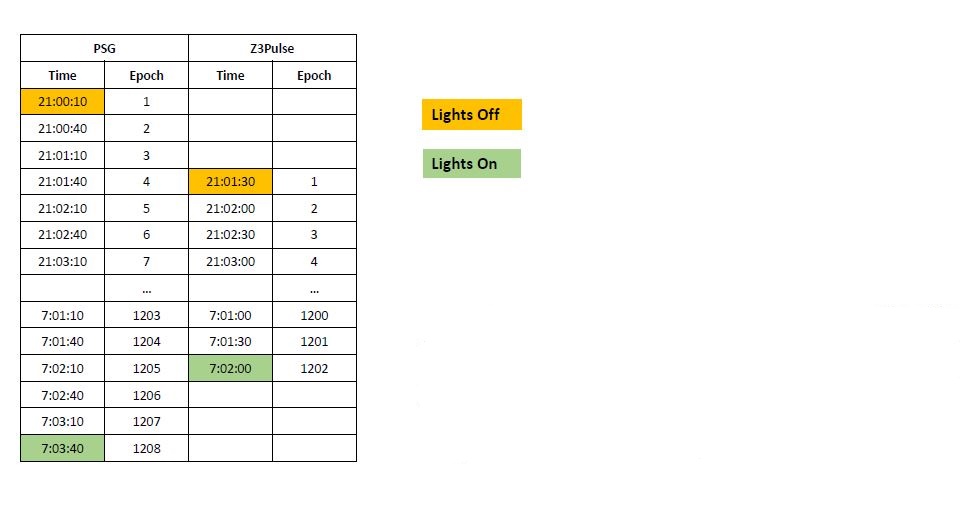
